# Modelling Road Accident Fatalities with Underdispersion and Zero-inflated Counts

**DOI:** 10.1101/2022.05.13.22275063

**Authors:** Teerawat Simmachan, Noppachai Wongsai, Sangdao Wongsai, Rattana Lerdsuwansri

## Abstract

**Background:** Thailand was rank second in the world in 2013 on the road accident fatality (RAF) rate, killing 36.2 of every 100,000 Thai peoples. In the past decade, during Songkran festival, the traditional Thai new year, the number of road traffic accidents (RTAs) was markedly higher than normal day life, but few studies have yet investigated this issue as the effect of festivity. The objective of this study was to investigate factors contributing to RAF using various count regression models.

**Methods:** Data of 20,229 accidents in 2015 were collected from the Department of Disaster Prevention and Mitigation, Thailand. Poisson, Conway–Maxwell–Poisson, and their Zero-Inflated versions were applied to analyze factors associated with the number of fatalities in an accident.

**Results:** The RAFs in Thailand follow a count distribution with underdispersion and excessive zeros which is rare. The best fitting model, the ZICMP regression model returns significant predictors (road characteristics, weather conditions, environmental conditions, and month) on the number of fatalities in an accident. The model consists of the count part encapsulating both non-excess zeros and death counts and the zero-part representing the considerable number of zeros during the festival months. The estimated proportion of the zero-part is 0.275 accounting for 5,563 non-fatal accidents. More specifically, the excessive number of no deaths can be explained by the month factor. The mean number of fatalities was lower in the festive periods than other months, with the highest in November.

**Conclusion:** For long, Thai authorities have put a lot of efforts and resources into improving road safety over the festival weeks, often they failed. This study indicates that people’s risk perception and public awareness of RAFs are mislead. Instead, nationwide road safety should have been announced by the authorities to raise the awareness of society towards everyday personal safety and the safety of others.

## Background

In 2013, Thailand ranked the second, after Libya, in the road accident fatalities (RAFs) for a survey of 180 countries in the world, with an estimated 36.2 deaths in road traffic accidents (RTAs) per 100,000 people [1]. In 2016, Thailand had an estimated rate of 32.7 RAFs and thus ranked the eighth of 175 countries and the first in Southeast Asia [2]. Using the RAFs prediction model as a function of registered vehicles per capita, the number of 30.68 deaths per 100,000 population in 2020 was estimated [3]. According to a 2020 report by the World Health Organization (WHO), these estimates of over 30 deaths per 100,000 people have remained virtually constant over the past decade, serving as evidence against the Thai government’s target for 2021 of 18 deaths as declared in the Road Safety Master Plan (2018-2021) [4]. Thailand seems unachievable neither the target of 10 deaths in the Decade of Action for Road Safety (2001-2020) nor the United Nations Sustainable Development Goal (SDG) 3 and its associated target 3.6 for reducing the number of global deaths and injuries from RTAs by 50% by 2020. This is also happening in other countries and thus the declaration of the second Decade of Action for Road Safety 2021-2030, with the target of reducing at least 50% of RAFs by 2030 [5].

Problems with the quality of the road safety databases are critical in Thailand. The country’s RAFs are particularly vulnerable to being underreported. In 2013, 14,056 deaths (20.98 deaths rate) were reported by the Office of Permanent Secretary, Ministry of Public Health. It was much lower than the estimated 24,237 deaths (36.2) by the WHO report. In response, the government emphasized its agenda on road safety issues. Lack of data integration was traced and first completed in 2016. The data have been managed and cleaned from three public sectors: (1) the Injury Surveillance System of Ministry of Public Health (MOPH) (death registration confirmed with medical certification of cause of death from hospitals); the Police Information System (POLIS) by the Royal Thai Police, Ministry of Interior; and (3) the E-Claim System by Road Accident Victims Protection Company Limited (all claim petitions and compensations related to RTAs national wide). With these collaborating efforts, the estimates by the WHO in 2016 (22,491 deaths) and the country’s report (21,745 deaths) were nearly synchronized. Later in 2018, the Injury Data Collaboration Center (IDCC) was set up and responsible for the management and maintenance of a national database on road safety [6]. The RAFs report for 2011-2020 over 76 provinces and the capital city of Bangkok can be accessible via the IDCC’s website, https://dip.ddc.moph.go.th/new/. It should be noted that the RAFs reported in Thailand is defined as unlimited period following crash, not by after crash immediately or within 30 days after traffic accidents [7].

Not only have the RAFs been focused over years, but also the numbers in a year have been critical, especially during the Songkran festival and New Year holidays. Fig 1 shows RAFs over the past decade (2011-2020). The RAFs for 2011-2018 have been more than 20,000 deaths per year and over 30 deaths per 100,000 inhabitants, representing about 58 deaths per day. During the long holiday of seven days, the RAFs rose to about 77 and 73 deaths per day for the Songkran festival and New Year holidays, respectively. In addition, real-time monitoring RAFs data over the seven-day deadly festivals show that more than 80% of victims died after crashes [8]. A lockdown policy for surveillance, detection and control of the COVID-19 epidemic in 2019-2020 seems to save life on the road. The lower number of cars on the roads proves that reducing traffic volume reduces RAFs. Other factors affecting the number of RTAs, RAFs and road accident injuries (RAIs) must be addressed to improve adequate preventive measures to overcome problems sustainably.

**Fig 1.**
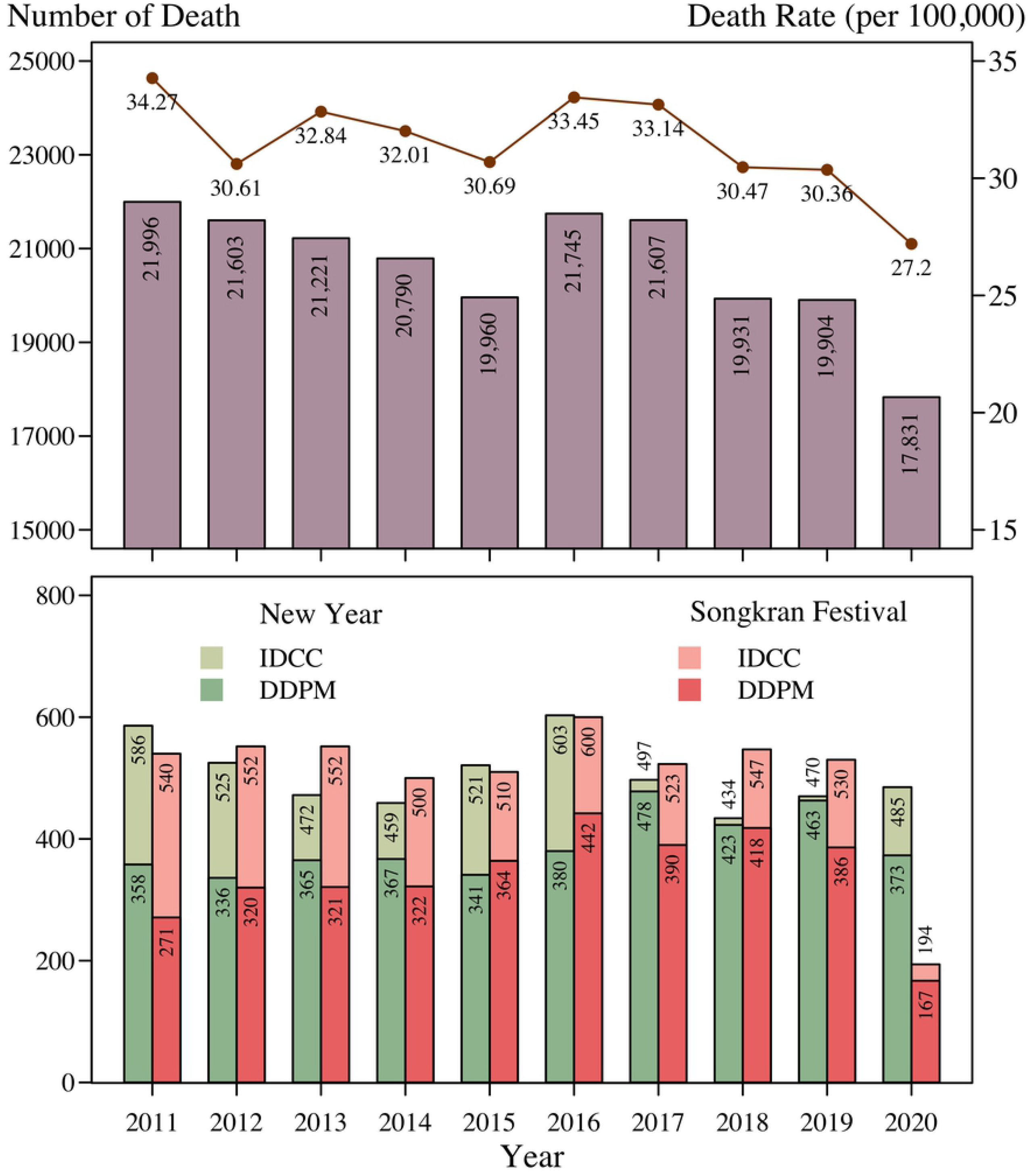
The number of deaths and death rate (per 100,00 population) on roads in Thailand for a decade of 2011-2020; overall (above) and during 7-day New Year Celebration and 7-day Songkran Festival (below) based on IDCC and DDPM reports.

Previous studies in Thailand have mainly focused on human behaviors and demographics rather than road and environment factors in explaining road safety incidences. Over the past two decades, RTAs, RAFs, and RAIs are dominated by motorcycles, young, drunk driving, and not wearing a helmet [4, 9-13], suggesting road safety campaigns and law enforcement failures. Few studies have recently considered road and environmental factors. The number of RTAs increased significantly with the higher volume of rainfall in the Southern and Northern provinces during 2012-2018 [14]. Such figures on Thai highways for 2011-2017 were affected by the length of the segment and average annual traffic volume [15]. Road type, road section and festive month were crucial factors relating to the number of injuries on roads [16].

Count regression models for road safety have been reported elsewhere. Differences in the distribution of accidents, deaths, and injuries could be attributed to the variation between roads and their surroundings. Several studies estimate the frequency of RTAs and/or RAFs by traditional Poisson regression [17], Negative Binomial (NB) regression [18-20], and Conway-Maxwell Poisson (CMP) regression [21-23], as well as estimate the count data with excessive zeroes by zero-inflated regression models [24-26]. However, predictive models for deaths and/or injuries are limited in Thailand road safety. The present study aims to fill this gap of research by using count regression models to investigate the distribution of RAFs in Thailand and its potential association with the road and environment at the accident location.

## Methods

### Data

Road traffic accident data were collected from the Department of Disaster Prevention and Mitigation (DDPM), Ministry of Interior for the year 2015, which was the most recent year of data availability at the time of this study. A total of 76 Disaster Prevention and Mitigation Provincial Offices is located across Thailand to handle disaster management at a local level. Each office has handled data collection, analysis, and reports on disaster damage and loss due to natural and man-made disasters including road traffic accidents.

In 2015, DDPM reported a total of 25,586 RTAs from emergency aid requests for crash accident victims at a provincial/district level. Completed data was available for 20,229 cases. The unit of analysis here is the individual RTA in a year of study, the dependent variable is the number of human deaths, and the independent variables are the road characteristics (class, surface, and section), weather condition, environmental condition, and month of the year. It should be noted that the national database on road safety supplies public information on the number of RTA, RTFs, types of vehicles, human demographics and behaviors, that are not of interest in the present study.

### Models

We applied a standard Poisson regression, CMP regression, Zero-Inflated Poisson (ZIP) regression and Zero-Inflated COM-Poisson (ZICMP) regression to model the number of fatalities in an accident for the year 2015, and its associations with road characteristics, weather conditions, environmental conditions, and month of the year.

### Poisson regression

The Poisson distribution is appropriate for a dependent variable *Y* taking nonnegative integer values 0, 1, 2, …. It can be used to model the number of occurrences of an event. In this study, *Y* is the number of deaths per accident. The probability mass function (p.m.f.), mean and variance of the Poisson distribution are shown in Table 1. Poisson regression has a limitation in its variance assumption. The expected death counts *λ*_*i*_ at the accident *i* (*i* = 1, 2,…, *n*) was calculated using 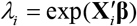 to ensure that *λ*_*i*_ > 0. 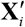 is the vector of covariates and **β** is the vector of estimable coefficients. Given the p.m.f., link function, and an assumption of independent observations, the log-likelihood function for observation *i* is given as

**Table 1.** Probability mass function, mean and variance for count distributions

| Models | p.m.f. | $E(Y)$ | $Var(Y)$ |
| --- | --- | --- | --- |
| Poisson | $\Pr(Y = y) = \frac{e^{-\lambda} \lambda^y}{y!}, y = 0, 1, 2, \dots, \lambda > 0$ | $\lambda$ | $\lambda$ |
| ZIP | $\Pr(Y = 0) = \pi + (1 - \pi)e^{-\lambda}, \lambda > 0, 0 \leq \pi \leq 1,$<br>$\Pr(Y = y) = (1 - \pi) \frac{e^{-\lambda} \lambda^y}{y!}, y = 1, 2, 3, \dots$ | $(1 - \pi)\lambda$ | $\lambda(1 - \pi)(1 + \pi\lambda)$ |
| CMP | $\Pr(Y = y) = \frac{\lambda^y}{(y!)^\nu Z(\lambda, \nu)}, y = 0, 1, 2, \dots,$<br>$Z(\lambda, \nu) = \sum_{j=0}^{\infty} \frac{\lambda^j}{(j!)^\nu}, \nu \geq 0, \lambda > 0$ | $\lambda \frac{\partial \log Z(\lambda, \nu)}{\partial \lambda} \approx \lambda^{1/\nu} - \frac{\nu - 1}{2\nu}$ | $\lambda \frac{\partial E(Y)}{\partial \log \lambda} \approx \frac{1}{\nu} \lambda^{1/\nu}$ |
| ZICMP | $\Pr(Y = 0) = \pi + (1 - \pi) \frac{1}{Z(\lambda, \nu)}, 0 \leq \pi \leq 1,$<br>$\Pr(Y = y) = (1 - \pi) \frac{\lambda^y}{(y!)^\nu Z(\lambda, \nu)}, y = 1, 2, 3, \dots,$<br>$Z(\lambda, \nu) = \sum_{j=0}^{\infty} \frac{\lambda^j}{(j!)^\nu}, \nu \geq 0, \lambda > 0$ | $(1 - \pi) \frac{1}{Z(\lambda, \nu)} \sum_{j=0}^{\infty} \frac{j \lambda^j}{(j!)^\nu}$ | $(1 - \pi) \frac{1}{Z(\lambda, \nu)} \sum_{j=0}^{\infty} \frac{j^2 \lambda^j}{(j!)^\nu} - E(Y)^2$ |

\* $Z(\lambda, \nu)$ is the normalizing constant.

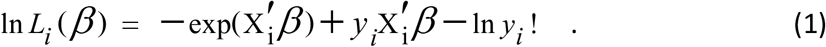

Summing over *n* observations, the log-likelihood function is

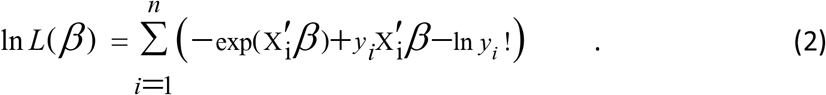

### Conway–Maxwell–Poisson regression

The CMP distribution is a generalization of the Poisson distribution serving to model both underdispersed and overdispersed data. This distribution was originally proposed by [27], but its implementation is contributed by [28-29]. The p.m.f., mean and variance are defined in Table 1. The CMP regression overcomes the restricted assumption of Poisson regression by defining an additional dispersion parameter, 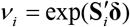. 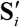 is the vector of covariates and **δ** is the vector of estimable coefficients. The likelihood function for observation *i* has the form

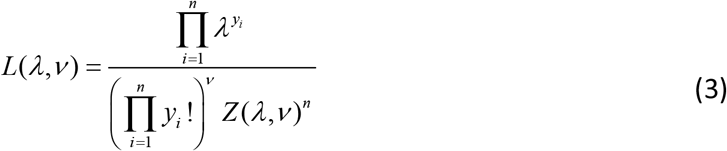

The log-likelihood function can be written as

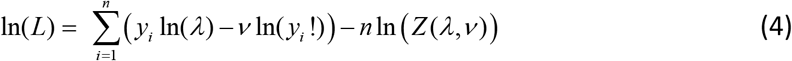

### Zero-inflated Poisson regression

The ZIP distribution is used to model count data that has an excess of zero counts. It assumes that with probability *π* the only possible observation is zero, and with probability1−*π*, a Poisson (*λ*) random variable is observed. The ZIP distribution is of the form

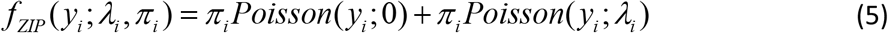

The resulting p.m.f., mean and variance are shown in Table 1.

The ZIP model can be generalized by a regression model, which allows both the Poisson parameter *λ*and the weight parameter *π* to vary. It has two parts: the Poisson count model predicting zeros and non-zero counts (count component of the model) and the logit model for predicting excess zeros, so-called always zeros (zero component of the model). 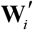 the vector of covariates and **γ** is the vector of estimable coefficients. The logit link is defined as follows:

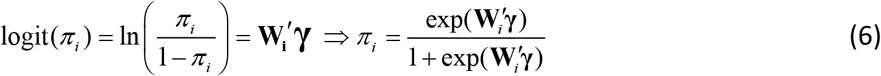

In the case that *π* is not a function of *λ*, the log-likelihood function for the ZIP regression is

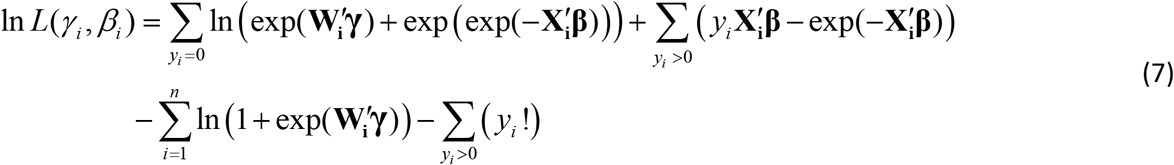

### Zero-inflated Conway–Maxwell–Poisson regression

A suggested model for underdispersed or overdispersed count data with excessive zeros is the ZICMP distribution proposed by [30]. This generalization of the ZIP distribution is a mixture of the degenerate at zero with probability *π* and a *CMP*(*λ,ν*) distribution with probability 1−*π*. Hence, the resulting p.m.f., mean and variance are shown in Table 1. The ZICMP regression allows both the CMP parameters (*λ,ν*) and the weight parameter *π*. It has two parts: the CMP count model and the logit model. The log-likelihood function for the ZICMP model is given by

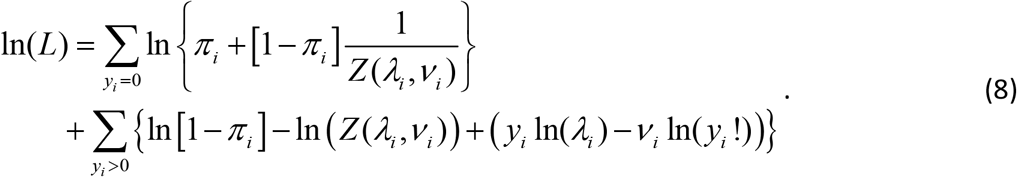

Maximum likelihood method was used to estimate coefficients for all four regression models used in this study.

### Dispersion test

A question arises if there is any evidence of dispersion. Firstly, exploratory data analysis is when a mean of death counts is greater (less) than its variance, an underdispersion (overdispersion) is observed. Additionally, the likelihood ratio test (LRT) can be performed for testing dispersion in CMP regression. According to the relationship between the mean and variance of the CMP distribution,

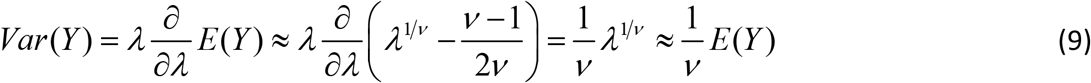

so that 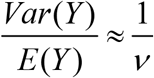. This yields the overdispersion and underdispersion for *ν* < 1 and *ν* > 1, respectively. Specifically, when *ν* = 1 the CMP model collapses to the Poisson model. Hence, the test of *H*_0_ :*ν* = 1 versus *H*_1_ :*ν* ≠ 1 can be interpreted as testing if the Poisson model is preferred over the CMP model. It should be noted that H_1_ does not specify the direction of data dispersion. In this case the LRT statistic is given by

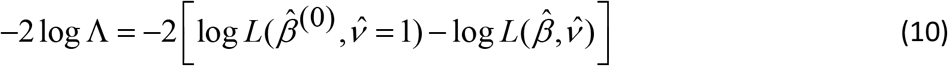

Where 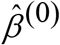 are the maximum likelihood estimates obtained under H_0_ (the Poisson estimates) and 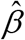 and 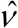 are the maximum likelihood estimates under H_1_ (the CMP model). The LRT statistic is asymptotically distributed as a chi-square with one degree of freedom [30].

Likewise, ZICMP can be reduced to ZIP if *ν* =1. The ratio of two log-likelihood functions of ZIP and ZICMP can be used as the following form 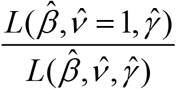. The LRT statistic

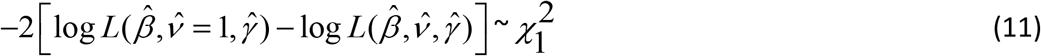

where 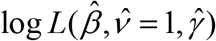 and 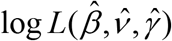 are the maximized log-likelihood under ZIP and ZICMP regression model, respectively.

### Zero-inflation test

The score test for zero inflation in a Poisson Distribution is a test for whether an extra proportion of zeros (always zeros part) is added to the common count part of the discrete Poisson distribution. Under the null hypothesis (*H*_0_ :*π* = 0), the score statistic has an asymptotic chi-squared distribution with 1 degree of freedom [31]. The score statistic is defined as

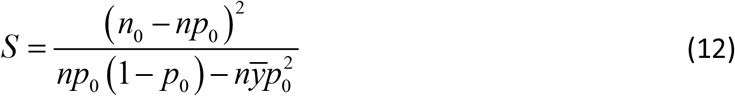

where *n* is the total number of observations, *n*_0_ is the number of zeros in the data, 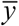 is the mean of death counts (the estimate of the Poisson parameter under the null hypothesis), and 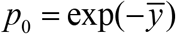.

Typically, CMP is a flexible, two-parameter distribution for overdispersed or underdispersed count data. A question addressing excessive number of zero counts under CMP is of interest leading to the ZICMP model. For testing a CMP model against a ZICMP alternative, the LRT test is determined by 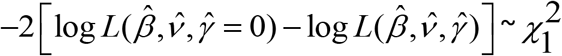 where 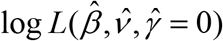 and 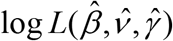 are log-likelihood values associated with CMP MLEs and ZICMP MLEs, respectively.

### Goodness of fit and model comparison

To assess the model adequacy, the generalized Pearson χ2 statistic is used and it is computed as follows.

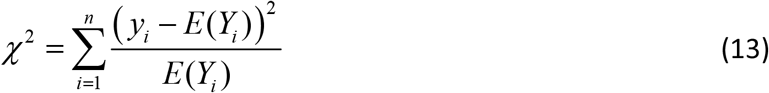

The numerator is the squared difference between the observed death count and the expected value of the death count, and the denominator is the expected value of the death count. In large samples, the distribution of this statistic is approximately chi-squared with n − k degrees of freedom, where n is the total number of observations and k is the number of estimated parameters including the intercept. The small value of the statistic tends to the decision to not reject the null hypothesis. If we fail to reject the null hypothesis, it shows that the model is adequate.

Assessment criteria used for model comparison are often based on several likelihood measures. Two of the most extensively used measures are log-likelihood value and Akaike Information Criteria (AIC). The model that provides a larger log-likelihood value is treated as a better model. The AIC is defined as 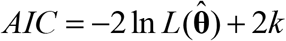, where 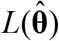 denotes the llikelihood function of a given fitted regression model and 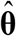is a vector of estimated parameters in the model. The smaller the AIC, the better the model.

### Software used

The results in this paper were obtained using R 4.1.0 [32] with four main packages for count models: MASS, pscl, COMPoissonReg, and vcdExtra. The standard Poisson model is described in a generalized linear model framework and implemented in R package MASS by the glm() function [33]. The zero-inflated extension of the Poisson distribution is provided by the zeroinfl() function in package pscl [34]. Currently, an implementation of count model for mean, dispersion, and zero-inflation is available in package COMPoissonReg by using glm.cmp() function [35]. The score test statistic is widely used for testing the null hypothesis that data does not characterize extra zeros for the Poisson distribution. The score test developed by [31] and has been implemented as a zero.test() in R under package vcdExtra by [36].

## Results

### Descriptive statistics

Distribution of death count is shown in Table 2 and Fig 2. The number of deaths per accident in 2015 ranges from 0 to 6, with many of the accidents reporting 0 or 1 death per crash. The proportion of death was highest for zero count (73%) and the second for one count (25.79%). Fatal accidents were about 27% of the total incidences (n=20,229), accounting for 6,109 deaths. 73% were non-fatal accidents in which at least one person was injured but no deaths occurred. The monthly average of RTAs was 465.7, 1,220.1, and 1,685.8 incidents for fatal, non-fatal, and total accidents, respectively. The number of RTAs was highest in April followed by January and December. These months celebrate important festivals in Thailand with a long holiday period of more than 5 days. However, it is noticeable that the fatalities per accident per month were lower in these festival months than other months. The highest proportion of 0.396 was seen in November, and the lowest of 0.165 was in April. A greater number of non-fatal injuries were seen during the celebrations. It plays a crucial role in excessive zero participation in the data.

**Table 2.**
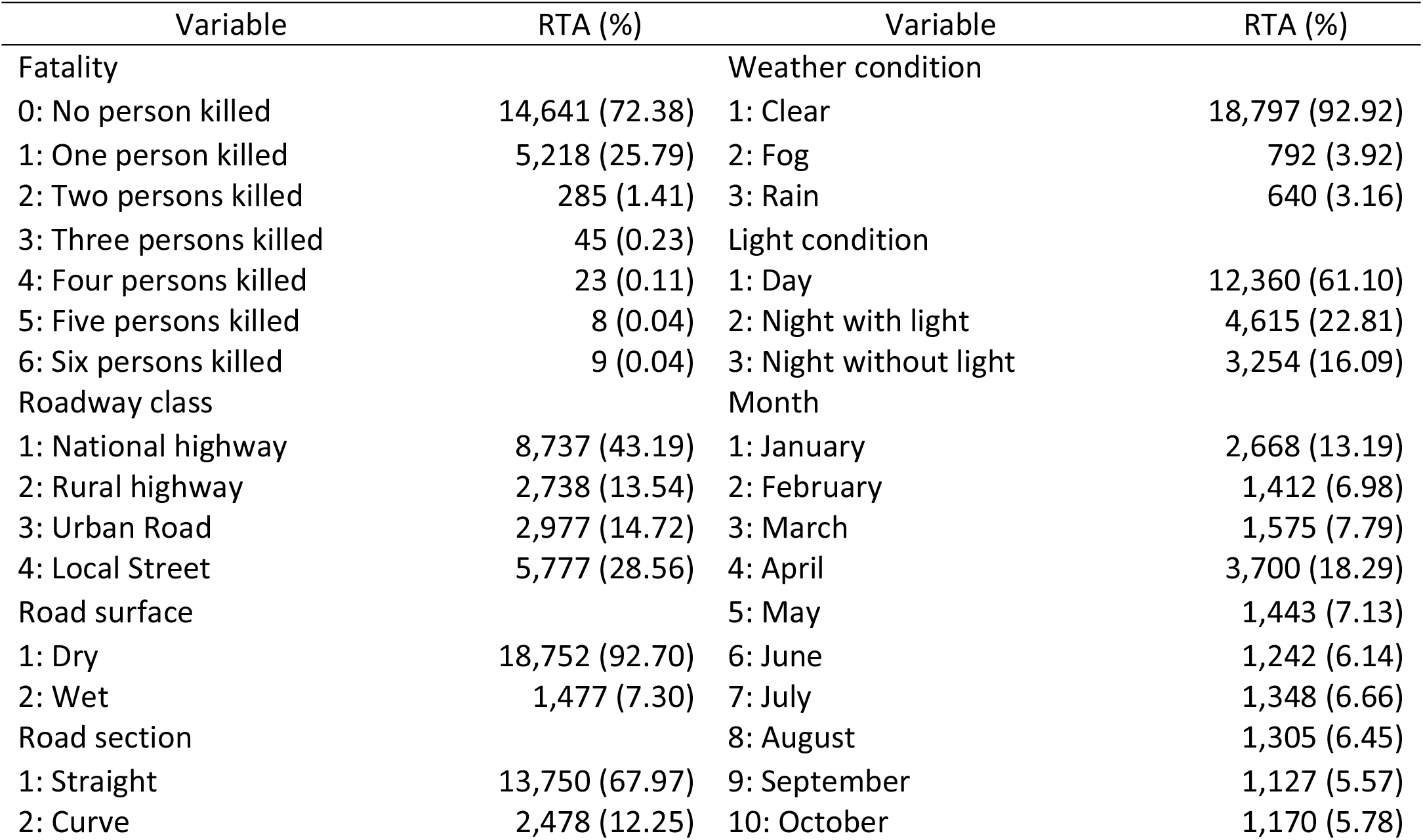
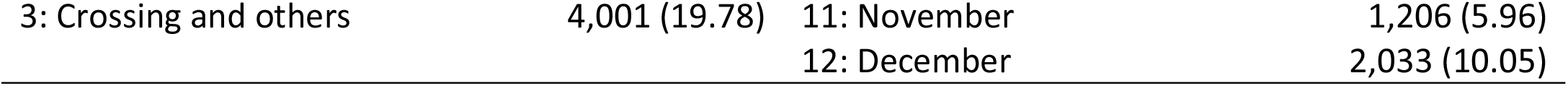
Variable description and the number of RTAs (percentage) in Thailand in 2015 (n=20,229) from the DDPM

**Fig 2.**
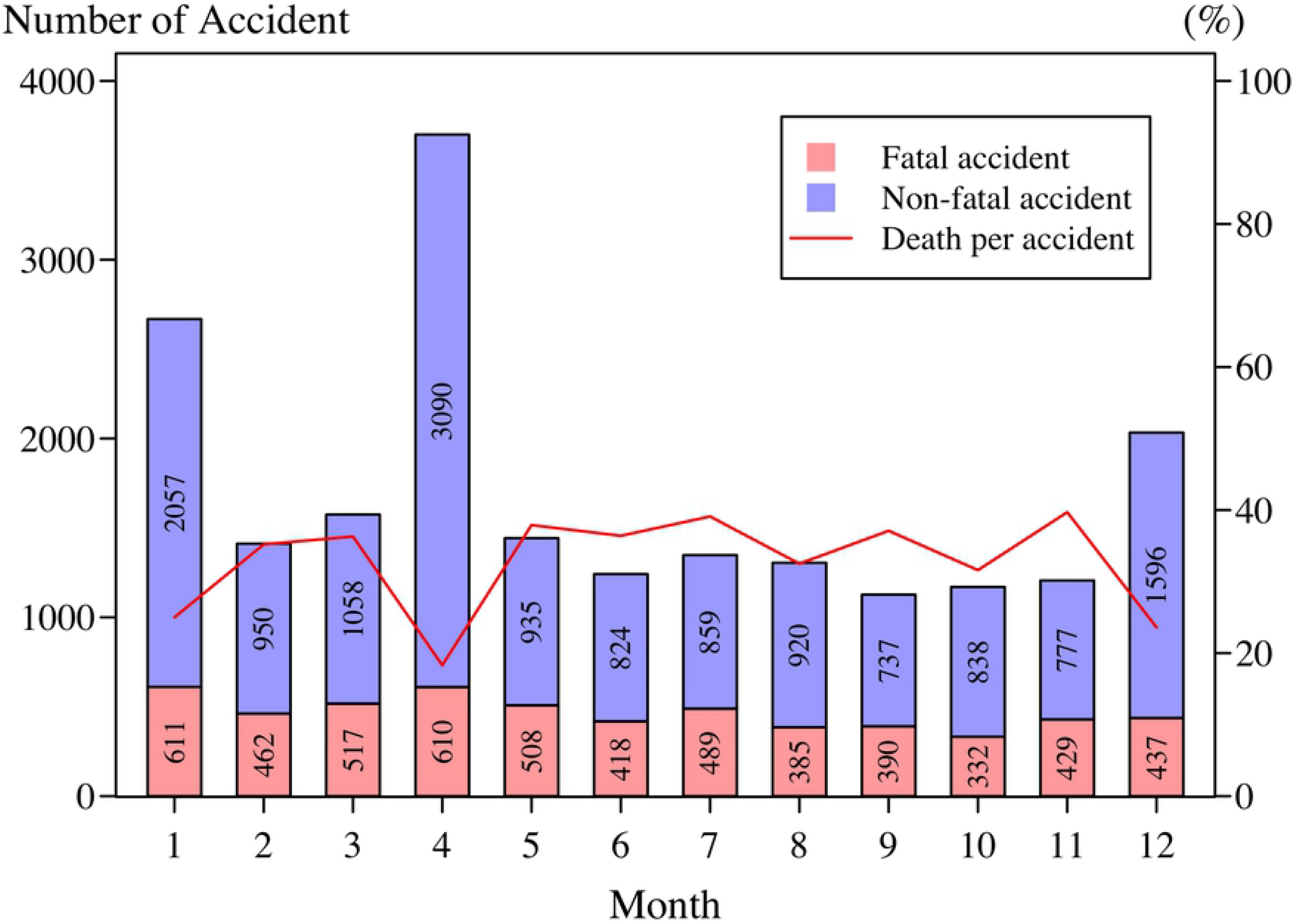
Statistics of the number of accidents and fatalities per accident by month in the year 2015.

Six explanatory variables were used in estimating road accident fatalities (Table 1). Many accidents occurred on national highways (43%) and rural highways (14%). These roads are constructed and maintained by the central government. The remaining accidents happened on urban roads (15%) and local streets (28%), building and maintenance by the local government administrative. The most common road geometry were straight section roads (68%), followed by pedestrian crossings (20%) and curves (12%). Dry surface roads were at elevated danger (93%) while roads with wet surface showed lower danger (7%). According to the light conditions, the accidents were reported in descending order during daylight (61%), at night with light on (23%), and at night without light (16%). Most accidents tended to occur in clear weather conditions (93%). During festival months, new year celebration (December: 10.1% and January: 13.2%), and Songkran holidays (April: 18.3%) presented higher frequencies of accidents, resulting in a sizeable number of fatalities and injuries.

### Fitting Fatality count distributions

Table 3 shows the results of fitting the death count data using four probability models. When fitting data to the Poisson distribution, the number of fatalities in a road accident, on average, was 0.302 (exp(−1.1974)) and its variance was 0.280. An estimated dispersion parameter in the Poisson distribution is thus less than one, indicating the count data with underdispersion. This situation was improved by fitting the CMP model to the data. The estimated dispersion parameter is 1.55 (exp(0.4384)), denoting underdispersion relative to the Poisson model (85.87, df =1, p-value < 0.0001). The expected number of deaths increases to 0.313 after adjusting for the dispersion (Table 1). The Pearson’s chi-squared statistic for the Poisson model is 19,239.34, while for the CMP model it is 5,812.78, a drop of 70%. Adding the dispersion parameter thus has much improved the model quality.

**Table 3.** Estimated parameters (standard error), log-likelihood value, AIC, Pearson’s Chi-square value for Poisson, CMP, ZIP, and ZICMP models

|  | Poisson | CMP | ZIP | ZICMP |
| --- | --- | --- | --- | --- |
| $\hat{\beta}$ | -1.1974<br>(0.0128) | -1.1030<br>(0.0163) | -1.1974<br>(0.0128) | -1.1030<br>(0.0163) |
| $\hat{\delta}$ | NA | 0.4384<br>(0.0404) | NA | 0.4384<br>(0.0404) |
| $\hat{\gamma}$ | NA | NA | -12.6700<br>(27.3600) | -14.4908<br>(42.3517) |
| Log-Likelihood | -13,872.42 | -13,829.48 | -13,872.42 | -13,829.48 |
| AIC | 27,746.84 | 27,662.97 | 27,748.84 | 27,664.97 |
| Pearson's Chi-square | 19,239.34 | 5,812.78 | 19,239.29 | 5,810.13 |
| #Parameters | 1 | 2 | 2 | 3 |

Note that statistically insignificant coefficients are presented in italics.

The CMP model simply decreases the variability in the Poisson distribution, but it does not necessarily accommodate the excess zeros. The score statistic gives evidence that the observed zeros exceed the zero bound of the Poisson distribution (177.69, df = 1, p-value < 0.0001). It is interesting that fitting the zero-inflated models to the data provided a different conclusion. The always-zero proportion parameter in the zero-inflation models was not statistically significant. Neither ZIP model nor ZICMP model captured the excess zeros in the data since the certain zero part was expected to be zero. Adding zero-inflation into a model did not improve the log-likelihood or reduce the AIC. It needs further explorations to fully explain the two processes of zeros in the RTAs data. In the next section, we will improve the fit by adding covariates to these models.

### Regression models with covariates

Count regression models were applied to investigate the relationship between the number of deaths and the covariates including road characteristics, weather conditions, light conditions, and month of the year. Results of coefficients and standard errors are shown in Table 4 and 95% confidence intervals in Fig 3. The standard Poisson regression and CMP regression were used to show the count-part modelling with and without an additional dispersion parameter, respectively. Then, the zero inflated models with ZIP and CMP regression were adopted for improving lack of model fitting in the zero-part modelling.

**Table 4.** Estimated parameters (standard error) from Poisson, CMP and zero-inflated regression models. Note that statistically insignificant factors are not presented

|  | Poisson | CMP | ZIP | ZICMP |
| --- | --- | --- | --- | --- |
|  | Count Part |  | Count Part |  |
| Roadway Class: baseline = National highway |  |  |  |  |
| Rural highway | -0.1715<br>(0.0356) | -0.2121<br>(0.0396) | -0.1606<br>(0.0361) | -0.2236<br>(0.0423) |
| Urban Road | -1.0079<br>(0.0500) | -1.1690<br>(0.0533) | -1.0013<br>(0.0504) | -1.2255<br>(0.0554) |
| Local street | -1.0136<br>(0.0378) | -1.1786<br>(0.0410) | -1.0206<br>(0.0382) | -1.2427<br>(0.0431) |
| Road Surface: baseline = Dry |  |  |  |  |
| Wet | -0.2528<br>(0.0722) | -0.2969<br>(0.0785) | -0.2389<br>(0.0730) | -0.2915<br>(0.0831) |
| Road Section: baseline = Straight |  |  |  |  |
| Curve | 0.1269<br>(0.0390) | 0.1555<br>(0.0433) | 0.1198<br>(0.0398) | 0.1777<br>(0.0470) |
| Crossing and<br>others | -0.1701<br>(0.0353) | -0.2047<br>(0.0384) | -0.1686<br>(0.0356) | -0.2258<br>(0.0404) |
| Weather: baseline = Clear |  |  |  |  |
| Fog | -0.1819<br>(0.0775) | -0.2190<br>(0.0842) | -0.1947<br>(0.0784) | -0.2287<br>(0.0895) |
| Rain | 0.2997<br>(0.0899) | 0.3599<br>(0.0998) | 0.3176<br>(0.0909) | 0.3941<br>(0.1063) |
| Light: baseline = Day |  |  |  |  |
| Night with<br>light | 0.1688<br>(0.0317) | 0.2034<br>(0.0350) | 0.1676<br>(0.0322) | 0.2170<br>(0.0374) |
| Night without<br>light | 0.4308<br>(0.0330) | 0.5301<br>(0.0373) | 0.42173<br>(0.0335) | 0.5518<br>(0.0401) |
| Month: baseline = January |  |  |  |  |
|  | Count Part |  | Zero Part |  |
| February | 0.3168<br>(0.0593) | 0.3814<br>(0.0653) |  |  |
| March | 0.3413<br>(0.0571) | 0.4126<br>(0.0629) |  |  |
| April | -0.3034<br>(0.0548) | -0.3481<br>(0.0588) | 1.2824<br>(0.2599) | 0.8632<br>(0.1480) |
| May | 0.3761<br>(0.0579) | 0.4548<br>(0.0639) |  |  |
| June | 0.3265<br>(0.0613) | 0.3940<br>(0.0676) |  |  |
| July | 0.4115<br>(0.0587) | 0.4984<br>(0.0649) |  |  |
| August | 0.2084<br>(0.0624) | 0.2472<br>(0.0683) |  | -1.1441<br>(0.4104) |
|  | Count Part |  | Zero Part |  |
| September | 0.3129<br>(0.0628) | 0.3770<br>(0.0693) |  |  |
| October | 0.1861<br>(0.0650) | 0.2209<br>(0.0712) |  | -0.7514<br>(0.3205) |
| November | 0.3954<br>(0.0601) | 0.4820<br>(0.0667) |  |  |
| December |  |  |  |  |
| $\hat{\beta}$ | -1.1002<br>(0.0427) | -0.9091<br>(0.0472) | -0.8269<br>(0.0236) | -0.4400<br>(0.0326) |
| $\hat{\gamma}$ | NA | NA | -1.7682<br>(0.2508) | -0.9714<br>(0.1350) |
| $\hat{\delta}$ | NA | 0.7381<br>(0.0316) | NA | 0.8654<br>(0.0297) |
| Log-Likelihood | -12950.99 | -12785.35 | -13017.66 | -12784.21 |
| AIC | 25946 | 25616.69 | 26081.31 | 25616.42 |
| Pearson's Chi-square | 17980.32 | 5294.07 | 17123.84 | 5260.38 |
| # Parameters | 22 | 23 | 23 | 24 |

**Fig 3.**
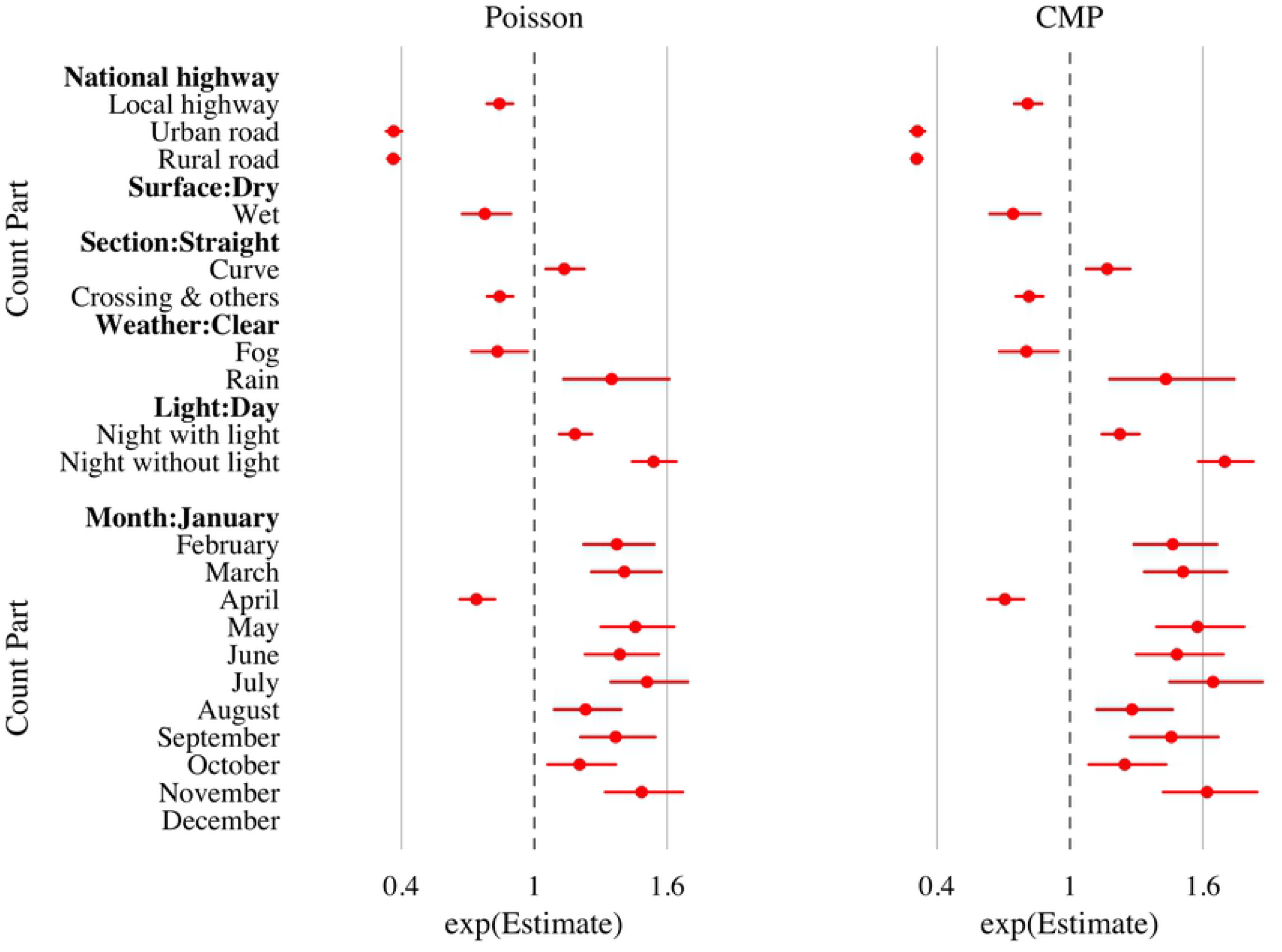
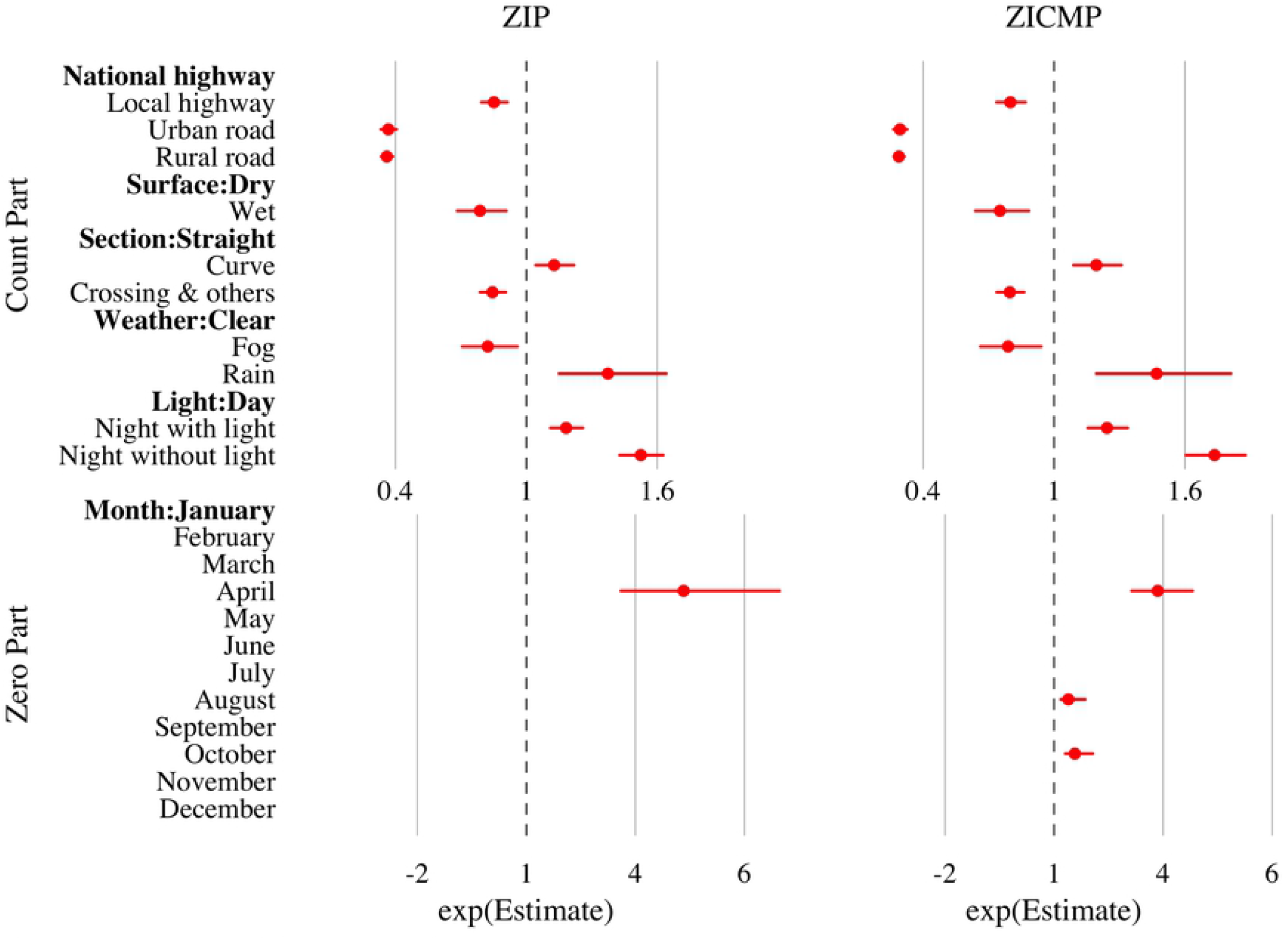
95% Confidence interval for statistically significant factors in Poisson, CMP and zero-inflated models

### Count part in Poisson and CMP regression

According to the log-likelihood, AIC, and Pearson’s chi-square value, the CMP regression outperformed the ordinary count regression. All six covariates were statistically significant predictors for the fatality distribution. Driving on the national highway or dry surface has a higher chance of having a higher risk fatality rate. On average, the number of deaths on the crossings is likely to decrease compared to that on the straight road sections. However, it is opposite when approaching curves. The positive light condition coefficient implies that driving during the daylight was much safer than driving at night with or without light on the streets. The RAFs were most common on sunny days. A higher risk was observed during rain but lower in foggy weather. Compared to the occurrence of RAFs in January, the fatality risk decreased in April, but increased in other months. For each predictor, the width of the 95% confidence intervals estimated from the CMP regression was wider than those from the Poisson regression. It is because of an adjustment for the underdispersion phenomenon.

### Count part and zero part in ZIP and ZICMP regression

The ZICMP model gives a better fit to the fatality frequency data than the ZIP model. Five risk factors were statistically significant for predicting the count part distribution while only the month of the year was the important covariate to extract the excessive zeros from the data. The estimated proportion of the always zeros was 0.275 (referred to Eq. (6)) suggesting that 5,563 (0.275 × 20229) observed zeros presented in the zero part. More specifically, the extra zeros reported the excessive number of no deaths that can be explained by the month factor. Using January as a baseline, the probability of zero deaths significantly increases in April but decreases in August and October. The number of zero deaths in other months did not differ from the first month of the year. These results confirm what we have seen in the exploratory data analysis that the death rate was lower during festival periods, as the unusually increased number of non-fatal accidents occurred. This information was not explicit when fitting the data with the Poisson or CMP regression. For the count part, the directions of associations between the response and the roadway class, surface, and section as well as lighting and weather conditions were the same as presented in the CMP regression result. But different magnitudes were clear since the ZICMP model allows the adjustment of the variability in the underdispersed data along with the zero inflation. The corresponding confidence intervals were estimated to reflect both phenomena. The expected count increased from 0.333 to 0.393 death per accident (referred to Table 1). Every ten accidents estimated four people are more likely to die.

## Discussion

### Model Selection

The most widely used model for analyzing count data is the Poisson model. There is a limit to the Poisson model as it assumes a unit variance-to-mean ratio. In road safety data, this restriction may be violated because the observed count may present a variance that is greater or smaller than expected, resulting in cases of overdispersion or underdispersion, respectively. The NB regression is often used to model RTAs, RAFs or RAIs with overdispersion, for example, the number of injury accidents on road bridges in Norway during 2010–2016 [18]; economic development and RAFs and RAIs with spatial panel data analysis in Thailand during 2012–2016 [37]; the numbers of human deaths per crash in the Oromia region of Ethiopia [19] and safety performance functions for urban intersections of Antwerp in Belgium [20].

The CMP regression is an alternative model to the NB regression and its advantage of handling both overdispersed and underdispersed count. The CMP is, however, not popular because of its complexity of estimating mean and dispersion parameters using iterative methods for nonlinear optimization and its limitation of computational tool availability. A generalized linear model based on the CMP distribution was developed by [38] and first applied to analyzing RTAs data with overdispersion by [21]. Two studies of crash data were from signalized four-legged intersections in Toronto, Ontario in 1995 and from rural four-lane divided and undivided highways in Texas. The results of a good-of-fit statistic and predictive performance were similar for the CMP and NB regression. Modeling RTAs, RAFs, or RAIs with underdispersed data is less reported. A study in 2010 by [22] shows that the CMP model for underdispersion provide better fit to RTAs than Poisson and gamma models for an analysis of 162 railway-highway crossings in South Korea between 1998 and 2002. A recent report in 2020 by [23] introduces the clustered longitudinal CMP with gamma random effects to model RTAs in Mauritius.

Excessive zero counts are highly skewed to the right and may cause a smaller conditional mean than a true mean value, resulting in overdispersion or underdispersion. Zero inflated version models of Poisson, NB, and CMP can be applied to overcome such problems. For example, the Poisson, NB, ZIP and ZINB were used to model of numbers of RAFs on the highest accident road F0050 in Malaysia [25] and in the Oromia region, Ethiopia [19]. Modeling zero inflated regression of Poisson and NB of the number of RTFs on the road F001 - Jalan Jb Air Hitam and on the road FT050 - Kluang-A/Hitam-B/Pahat, Malaysia was presented by [24] and [26], respectively. The results suggest that the zero inflated version models offered better statistical performance than the traditional models. Recently, the ZICMP distribution and regression along with their computational “COMPoissonReg” package in R were developed by [30]. The ZICMP can account for the excess zeros and the data with overdispersion or underdispersion. The model performance was compared to the ZIP and ZINB in a study of educational data with overdispersion. It may be as expected that the ZICMP and ZINB provide a similar performance in capturing extra zeros and overdispersion and surpassing the ZIP. To our best knowledge, it is not yet used in road safety analysis. In this study, we fit the Poisson, CMP, ZIP, and ZICMP regression models to RAFs occurred in Thailand in 2015. It is attractive since excess zeros and underdispersion are rarely seen in road safety data. Thus, the ZICMP model performs the best. The ZICMP slightly outperformed the CMP as their difference in log-likelihood values is very small and maybe negligible. Such that two distinct aspects are clear. Having a lot of zeros does not necessarily mean that a zero inflated model is needed. It is a matter of what issues of concern need to be addressed and the possibility of finding associated factors. The CMP gives insight knowledge about death counts per crash if no extra zeros. It is common that when an accident occurs, a driver or passengers may be injured or die. The ZICMP accounts for excessive zeros, suggesting an unusually increasing number of no deaths on roads. This can be viewed as unexpected non-fatal accidents. Extraction of such information requires identifying associated covariates.

### Month, road and environmental effects

The month of the year plays a vital role in extracting different messages from our underdispersed fatality data. In case of death counts, the count part in CMP addresses the lower number of fatalities in April and the larger number of deaths in other months, compared to January. For the extra zeros, the zero part in ZICMP captures excess zero deaths in April but lower figures in August and October, compared to January. However, the month did not affect the death counts in the count part in the ZICMP model. It reflects that different sets of variables are needed to show different meanings from the data. Our result serves the purpose of the model formulation. These two generalized linear mixed models allow different sets of relationship between the average, dispersion or zero structure and its associated covariates [30].

Two key findings are highlighted in April: a lower number of death count and a greater number of excess zero death on roads. These reflect the fact in this festival month. For centuries, April is celebrated annually as the traditional Thai New Year, known as the Songkran festival (the period of 11-17 April every year). It is rich in culture and tradition of water blessing ceremonies for prosperity and wellness as well as the world-renowned water parties for ritual cleansing and a fresh start. An unacceptable drawback is the time of the highest crash frequencies of the year, called the “seven deadly days”. Intensive road traffic occurs at the beginning and end of the long holiday when there is an excess of public transport and private vehicles on part of the roadway connecting the provinces. Since the rail public transit and railway network in Thailand are not efficiently served, public buses need to be double or even triple the number of services to accommodate the most prominent human migration of the year. The traffic flow rate on the road is low, causing traffic jams and accidents. Road accidents may have caused more congestion, especially on interconnected highways and national highways.

For decades, the government has put a lot of time, budget and effort into road safety campaigns to raise awareness and change attitude and behavior, especially during festivals. But so far, all earlier campaigns have shown no reductions [9,11-13]. A new insight from our study showed that the number of deaths was not quite different between the months but the number of nonfatal accidents. Interestingly, our findings provide contradiction in Thai people’s risk belief and public awareness of road accident fatalities. The advertising campaigns emphasize the death counts, not mentioning the accident counts. They shout as if the number of deaths on roads during festivals were dramatically higher than usual. This is deceptive. Accidents related to injury and property damage are also harmful to social and economic society. Disability after RAIs is a massive health problem and causes economic consequences of a loss of gross domestic product in the country by 3% - 6% or 273-545 million Baht (US$ 8.3-16.5 million, based on the exchange rate of 33 Baht per US$ 1) [37]. We therefore urge and recommend that the authorities implement adequate road safety measures for road users daily, not just for a specific period.

Most of the literature has in common that factors affecting RTAs and RAFs are roads and the environment. In this regard, the present study gave comparable results. The correlations between traffic accidents and environmental factors like population and road conditions were studied by [39]. Female, the presence of alcohol and high degree curve would increase the likelihood of severe traffic accidents [40]. Human behavior and demographics are also known as the factors associated with road safety [41], and thus may be considered in further studies in Thailand.

## Conclusion

In this study, the CMP regression and its zero-inflation version ZICMP regression were delivered to handle an underdispersion problem and excessive zero death counts for the road traffic fatalities in Thailand. Neither Poisson regression nor ZIP regression is proper for modelling this type of count response. Since the underdispersion is not accounted for, the classical models produce a lower standard error of estimated parameters than expected. We highlighted that having many zeros does not necessarily mean that a zero inflated model is needed. It is a matter of what issues of concern need to be addressed and the possibility of finding associated factors for capturing such zero-inflation issues. A key finding is that the month of the year has a power to figure out the difference between the death counts and the extra zero death counts, referred to as uncommon nonfatal accidents in our case. Using January as a baseline, the ZICMP regression revealed the largest number of nonfatal accidents in April when people celebrated the Thai traditional New Year, Songkran festival. This figure was not elucidated by using Poisson or CMP regression. The CMP regression shows the lower fatality rate during this festival month since the number of deaths was not altered much throughout the year, but the number of accidents was considerably higher in April than other months. Other covariates including road (class, surface, and section) and environment (weather and light conditions) were significant predictors of road traffic fatalities in Thailand as commonly reported elsewhere.

The underdispersion problem in road accident data is a rare case. However, it was presented in the road traffic fatalities in Thailand. In addition to the CMP model, the geometric model may serve as an alternative model to the familiar Poisson models. The advantage of the geometric distribution is that it is a mixture of a Poisson with an exponential distribution, thus it incorporates some form of heterogeneity. We leave this point as future work. Additionally, human behavior, vehicles, roads and environment interact with each other in the process of accidents. A possible enhancement of road safety database in which various authorities involved to supplying such data is key to effective road safety measures to reduce road accidents, injuries and fatalities, not only in Thailand but worldwide.

## Data Availability

The data underlying the results presented in the study are available from https://data.go.th/

## Acknowledgement

The authors acknowledge with thanks the availability of data coming from the Department of Disaster Prevention and Mitigation. The authors also thank the editor along with anonymous reviewers for their useful comments. This research was supported by Thammasat Postdoctoral Fellowship [grant number TUPD7/2564].

## Author Contributions

**Conceptualization:** Teerawat Simmachan.

**Data curation:** Teerawat Simmachan, Noppachai Wongsai, Sangdao Wongsai, Rattana Lerdsuwansri.

**Formal analysis:** Teerawat Simmachan, Noppachai Wongsai, Sangdao Wongsai, Rattana Lerdsuwansri.

**Investigation:** Noppachai Wongsai.

**Methodology:** Teerawat Simmachan, Sangdao Wongsai, Rattana Lerdsuwansri.

**Resources:** Sangdao Wongsai.

**Software:** Noppachai Wongsai.

**Supervision:** Sangdao Wongsai.

**Validation:** Sangdao Wongsai.

**Writing – original draft:** Teerawat Simmachan, Noppachai Wongsai, Sangdao Wongsai, Rattana Lerdsuwansri.

**Writing – review & editing:** Teerawat Simmachan, Noppachai Wongsai, Sangdao Wongsai, Rattana Lerdsuwansri.

